# Elevated Blood Glucose in COVID-19 Patients: An explorative Study

**DOI:** 10.1101/2024.11.15.24317345

**Authors:** Maryam Bisher, Ahlam Thamer, Shaheen Shahata, Ahmed Atia

## Abstract

**Background:** Since the emergence of the coronavirus pandemic in 2019, scientific studies have not stopped to understand the mechanism of several disorders that occurred in patients who were infected with the COVID-19 virus, among these disorders, the most important of which is high blood glucose because of its impact on human health. This study aimed to identify the association between blood glucose concentration and some biochemical variables that lead to high blood glucose in people infected with Covid-19 virus.

**Methods:** This study was conducted on 100 samples of serum of people who were confirmed to be infected with the COVID-19 virus using polymerase chain reaction (PCR) technology. Samples were collected from the Broccoli Isolation Center, the Respiratory Clinic, and the Sebha Medical Center in Sebha city, Libya. The concentration of glucose, insulin, cortisol, triglycerides, cholesterol, CRP and liver enzyme’s activity was measured.

**Findings:** The results showed that 60% of the patients had an increase in blood glucose concentration, and 40% of the patients had a normal glucose concentration. The levels of insulin, cortisol, CRP, triglycerides, and liver enzyme’s activity were compared between the two groups, and the results showed an increase in the mean concentration of cortisol, triglycerides, CRP and liver enzymes in the group of patients with high glucose concentration compared to the group of patients with a normal concentration of glucose, and the statistical analysis using the t-test showed that there were significant differences between the means (P = 0.000), while the mean insulin concentration was lower in the group of patients with elevated glucose level. The results also showed positive correlation between glucose and cortisol, triglycerides, CRP concentration and GPT, GOT activity.

**Interpretation:** Patients infected with the COVID-19 virus had elevated blood glucose concentration associated with decrease in their insulin hormone concentration. In addition, high concentration of biochemical variables that contribute to high blood glucose in patients infected with the COVID-19 virus were noticed.

## Introduction

Coronavirus disease 2019, or COVID-19, also known as acute respiratory disease associated with the novel coronavirus 2019, is a disease that affects humans after exposure to a coronavirus or rectal coronavirinate (Orthocoronavirinae), a subfamily of single-chain RNA viruses that infect mammals and birds.^1^ Since the beginning of the COVID-19 pandemic, studies have shown that diabetes is one of the main comorbidities associated with the development of COVID-19 symptom severity and mortality, ^2^ adults with diabetes are at greater risk of acute respiratory distress syndrome (ARDS), pneumonia and excessive responses to uncontrolled inflammation and hyper coagulation. ^3^ Recently, a new hypothesis has emerged that assumes a bidirectional relationship between diabetes and COVID-19, so that not only does the presence of diabetes increase the likelihood of developing complications of COVID-19, but the virus may lead to the development of diabetes. ^4^ Many studies conducted on the relationship of the COVID-19 virus to diabetes ask: have all people with COVID-19 developed diabetes or not? Was this infection due to the virus or were patients who showed high blood glucose had diabetes before contracting the virus but were only discovered after the person was exposed to the virus? And what are the reasons that led to high blood glucose, ^5^ and other studies have reported that high glucose in the blood of patients infected with the COVID-19 virus has severe complications for those infected, as those with high blood glucose (whether previously with diabetes or those who have recently had diabetes) suffered from the worst results in the development of COVID-19 disease compared to patients who did not have high blood glucose and those without diabetes. ^6^ After the appearance of these complications in patients with the COVID-19 virus, and after the researchers confirmed the occurrence of a rise in blood glucose in patients, research began to search for the causes of high glucose in the blood after infection with the Covid-19 virus. What are the factors that led to this? Can all patients develop diabetes while they are infected with the virus? Does the disease continue with them even after they recover from the COVID-19 virus, and for this reason many hypotheses have been put forward to explain the mechanism of diabetes in patients, including:1-COVID-19 attacks multiple organs in the body, including the pancreas? ^7^

Previous studies have found that the receptors of this virus are present in the endocrine tissues of the pancreas, which led to damage to the cells of the islets of Langerhans (insulin-producing area), the entry of the virus into these tissues leads to complex imbalances in glucose metabolism, impaired insulin secretion leads to high blood glucose and diabetic ketoacidosis (DKA), which may be the cause of type I diabetes, and researchers noted that most cases of diabetes that arose during hospitalization disappeared within three years. ^8^ Some prevalent treatments for the COVID-19 virus are the use of glucocorticoids, a class of steroid hormones, which work on carbohydrate metabolism and insulin hormone antagonists, this treatment can cause a significant variation in blood glucose, this may be the cause of type II diabetes. ^9^ 3-The relationship between the COVID-19 virus and diabetes may be related to inflammation, as previous studies indicate a relationship between diabetes and the occurrence of inflammation in the body, as inflammation leads to an immune response that carries out a series of reactions that result in insulin resistance, which may lead to type II diabetes. ^10^ 4-High body mass index, overweight and obesity were also one of the hypotheses developed to explain the cause of high blood glucose and the new diabetes caused by COVID-19, as obesity complicates the body’s glucose metabolism, ^11^ and high blood glucose is closely related to obesity, which plays an important role in disease severity and mortality, ^12^ some studies conducted on COVID-19 patients have recorded the presence of cases of obesity in patient profiles, associated with high glucose during their period of infection with the virus, and a reference study that included a number of studies around the world indicated that obesity may also increase the risk of infection with the Covid-19 virus, meaning that the relationship between obesity and the Covid-19 virus is a bilateral mutual relationship, that is, obesity is a cause of high glucose in COVID-19 patients, and on the other hand, obesity is One of the factors that increase the risk of infection with the COVID-19 virus, ^13^ reports issued by the World Health Organization have indicated that during the quarantine period, which was one of the measures to prevent infection with the virus during its spread, which spread almost all countries of the world, has led to a decrease in the physical activity of people with the presence of unhealthy eating, all factors led to weight gain and changes in insulin resistance, which led to an increase in the likelihood of The onset of diabetes.^14^ This study aims to find out the relationship between the concentration of blood glucose and some biochemical variables that lead to high blood glucose in people infected with COVID-19.

## Methods

### Study design and sample

100 samples of people infected with the COVID-19 were collected from isolation centers in the city of Sebha (Libya), including the Barcoli isolation center, the respiratory clinic and the Sebha Medical Center, where the mean age of the infected was 57.07± 20.87 years, divided into 36 females and 64 males, their diagnosis of COVID-19 was confirmed using PCR technology, where the test result was positive for all patients from whom samples were collected. These samples were collected in tubes containing the lithium heparin anticoagulant, on which all biochemical tests can be measured without any interference and giving false results.

### Procedures

The measurement was done using the Cobas Intigra 400 plus device, which is a fully automatic and computerized chemical analyzer device. The hormones were measured on the Cobas e 411, a fully automated analyzer using patented ElectroChemiluminescence (ECL) immunoassay analysis technology.

### Homeostatic Model Assessment of Insulin Resistance (HOMA-IR) and beta cell function (HOMA-*B*)

The homeostasis model assessment of insulin resistance (HOMA-IR) and beta cell function (HOMA-□) constitute a method for assessing beta cell function and insulin resistance from glucose and insulin concentrations. To calculate the value of insulin resistance and B function, the following equations is used:

HOMA-IR = fasting insulin (µU/ml) × fasting plasma glucose (mg/dl)]/405 ^15^ HOMA-□% = fasting insulin (µU/ml) × 360 / [fasting plasma glucose (mg/dl) – 63] ^15^

### Statistical analysis

The data were statistically analyzed using Minitab version16, we used a 2-sample t-test and confidence interval to analyses variance between groups and person correlation between variables at a significant level of ≤0.05.

## Results

### Distribution of incidence based on age

Following the collection of samples, individuals infected with the Covid-19 virus were categorized according to their age groups, aiming to identify the most affected age group. The analysis revealed that approximately seven (7%) of the infections were among adolescents, aged 18 to 24, while 50(50%) were observed in adults aged 25 to 64. Furthermore, 43 (43%) of the patients were elderly, aged 65 and above, as depicted in (Figure 1).

**Figure 1.**
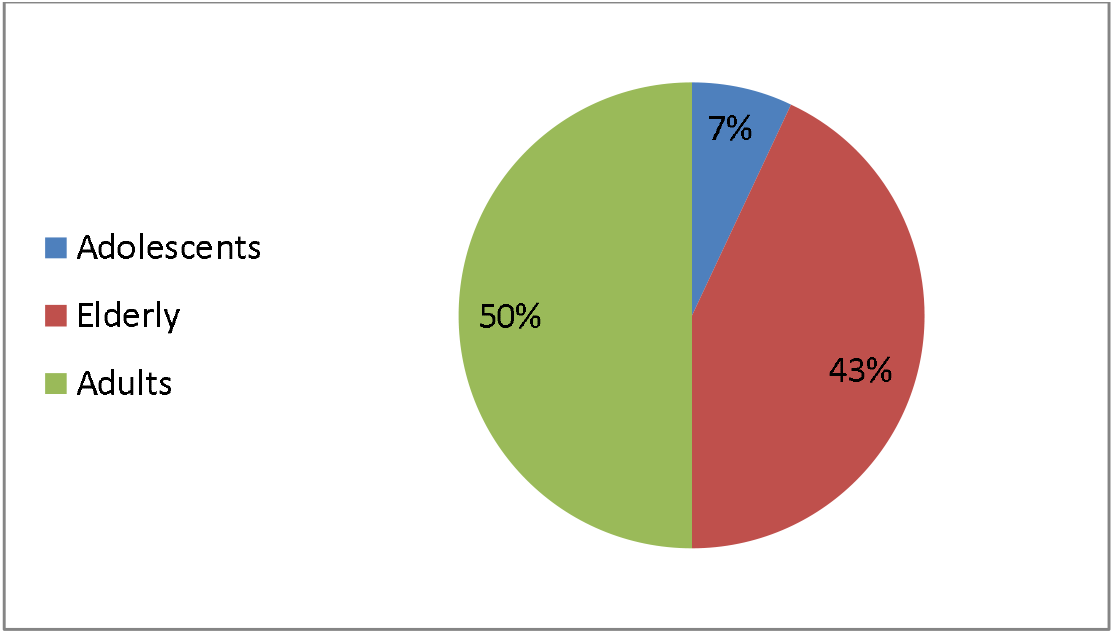
Distribution of incidence based on age.

### Comparison of the concentration of biochemical variables in patients after dividing them based on blood glucose concentration

Patients were divided into two groups based on blood glucose levels: a group with high glucose levels consisting of 60 individuals, and a group with normal glucose levels consisting of 40 individuals. Mean concentrations of insulin, cortisol, C-reactive protein, triglycerides, cholesterol, and liver enzyme activity were analyzed in both groups. The results indicated that the mean cortisol concentration in the group of patients with high glucose levels was significantly higher than that in the group with normal glucose levels, as confirmed by statistical analysis using the T-test (*P* = 0.000). Furthermore, the mean insulin concentration was lower in the group of patients *with* high glucose levels compared to the other group, with a significant difference between the two means (*P* = 0.000). Similarly, the mean concentrations of C-reactive protein, triglycerides, and cholesterol were higher in the group of patients with high glucose levels compared to the other group, with significant differences in the means for these variables (*P* = 0.000). In addition, the results showed that the mean activity of liver enzymes was higher in the group of patients with high glucose concentration compared to the other group, with significant differences in these means within the two groups, except for the ALP enzyme, where statistical analysis did not indicate significant differences in the mean activity of this enzyme between the two groups (Table 1).

**Table 1.** Comparison of the concentration of biochemical variables in patients after dividing them based on blood glucose concentration.

| Variables | Patients with normal glucose (n=40) | Patients with high glucose (n=60) | *P value* |
| --- | --- | --- | --- |
| Glucose(mg\dl) | 87.7(11.8) | 200.9(79.3) | 0.000* |
| Insulin(μU/mL) | 11.7 (5.8) | 3.4± 1.11 | 0.000* |
| Cortisol (nmol/L) | 255.1 (70.2) | 592 ± 222 | 0.000* |
| CRP (mg\dl) | 22.9 (12.1) | 66.6 ± 30.2 | 0.000* |
| TG (mg\dl) | 134.3 (40.2) | 257 ± 130 | 0.000* |
| CHOL(U\L) | 130.8 (39.3) | 319 ±78.7 | 0.000* |
| GOT(U\L) | 30.7 (12.1) | 106.5 ± 79.9 | 0.000* |
| GPT(U\L) | 39.8 (15.4) | 145 ± 30.7 | 0.000* |
| GGT(U\L) | 21.5 (11.9) | 54.6 ± 16.4 | 0.000* |
| ALP(U\L) | 144.8 (20.6) | 171 ± 18.2 | 0.113 |

*Data are mean (SD), n (%), \*= Significant*

### The correlation coefficient between glucose and other variables in the group of patients with high glucose concentration

When examining the correlation coefficient between glucose concentration and other variables in patients with hyperglycemia, the results indicated a significant strong negative correlation and a significant strong positive correlation between glucose concentration and insulin and cortisol levels (r = - 0.63, *p* = 0.000, r = 0.61, *p* = 0.000) respectively. In addition, there was a noteworthy positive correlation with C-reactive protein concentration (r = 0.43, *p* = 0.000), and a weak positive correlation with both cholesterol and triglyceride concentrations (r = 0.29, *p* = 0.002, r = 0.39, *p* = 0.002) respectively. Regarding liver enzymes, correlation analysis showed a weak positive correlation between glucose concentration and GOT and GPT enzyme activities (r = 0.3, *p* = 0.02, r = 0.35, *p* = 0.006) respectively, while there was no significant correlation between glucose concentration and GGT, ALP activities (r = 0.18, *p* = 0.15, r = 0.2, *p* = 0.19) respectively (Table 2). When testing the correlation coefficient between glucose concentration and other variables in the group of patients whose glucose was normal, the results showed that there was no association between them (P>0.05).

**Table 2:**
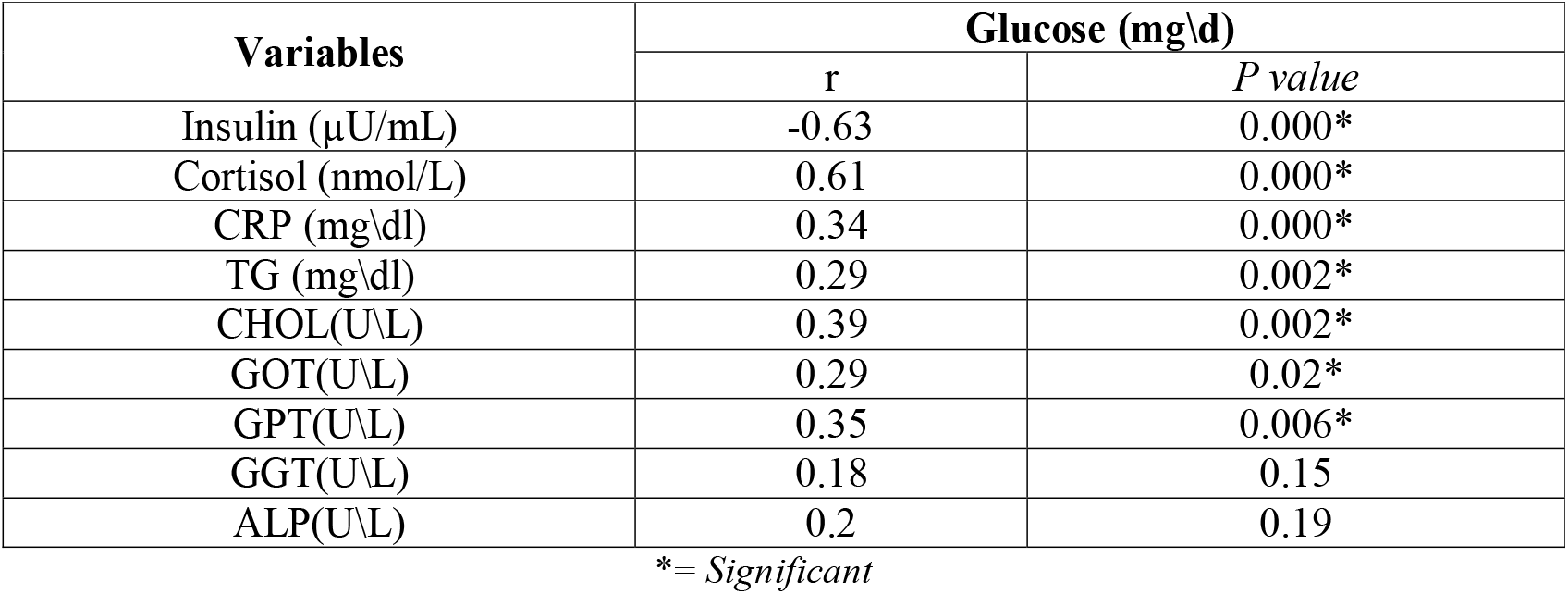
The correlation coefficient between glucose and other variables in the group of patients with high glucose concentration.

| Variables | Glucose (mg\dl) |  |
| --- | --- | --- |
|  | r | P value |
| Insulin (μU/mL) | -0.63 | 0.000* |
| Cortisol (nmol/L) | 0.61 | 0.000* |
| CRP (mg\dl) | 0.34 | 0.000* |
| TG (mg\dl) | 0.29 | 0.002* |
| CHOL(U\L) | 0.39 | 0.002* |
| GOT(U\L) | 0.29 | 0.02* |
| GPT(U\L) | 0.35 | 0.006* |
| GGT(U\L) | 0.18 | 0.15 |
| ALP(U\L) | 0.2 | 0.19 |

\*= Significant

### HOMA-IR calculation

The HOMA-IR value was calculated for the group of patients whose glucose concentration was normal, and the results showed that their mean insulin resistance value was 2.54±1.27 which is a high value when compared to its reference values, which means that this group has high insulin resistance (Table 3). When testing the correlation coefficient between HOMA-IR and concentration of insulin and between glucose and insulin concentration, it was found that there was a significant, strong positive correlation (r = 0.9, *p* = 0.000), and a significant negative correlation (r = - 0.3, *p* = 0.04) respectively.

**Table 3.** Comparison of HOMA-IR and HOMA-□ values in both groups

| Variables | Patients with high glucose (n=60) | Patients with normal glucose (n=40) | P value |
| --- | --- | --- | --- |
| HOMA-IR | 2.55 ± 1.27 | 1.25 ± 1.15 | 0.000* |
| HOMA-β | 352.46 ± 124 | 18.26 ± 17.55 | 0.000* |

\* = Significant

### HOMA-□ calculation

The results showed that mean of HOMA-□ in patients with high glucose concentration (accompanied by low insulin concentration was less than the normal level, and patients with normal glucose concentration (accompanied by high insulin concentration) was mean of HOMA-□ high. Compared to another group (*p* = 0.00) (Table 3). When measuring the correlation coefficient between HOMA-□ and insulin concentration in both groups, we found that there was a strong positive correlation between them (r = 0.96 *p* = 0.000, r = 0.51, *p* = 0.002) respectively.

## Discussion

One of the reasons that led to deaths during the period of the emergence of the COVID-19 is high blood sugar, and opinions differed about the reason for the increase in diabetes in those infected patients. Numerous studies had reported that the frequent occurrence of high blood sugar levels in many individuals were linked to large number of deaths. This confirms its status as a critical and severe complication of the disease. ^16^ People with diabetes are particularly susceptible to respiratory issues, breathing difficulties, and weakened immune responses, as evidenced by data from Canada in September 2020, aligning with findings from other nations. Studies have highlighted the heightened risk of severe COVID-19 infection in individuals with diabetes, ^17^ resulting in a higher mortality rate compared to non-diabetic COVID-19 patients. Moreover, it is noteworthy that even non-diabetic COVID-19 patients had presented with elevated blood sugar levels. ^18^ The impact of COVID-19 on blood sugar levels raises questions about whether the virus induces dysregulations in glucose levels by compromising insulin secretion, potentially leading to the long-term development of diabetes. Another aspect to consider is whether irregularities occur as a result of infection-related factors contributing to increased blood glucose levels. Various theories have been proposed to elucidate the connection between COVID-19 and blood glucose elevation. One theory suggests that the virus may cause damage to the pancreatic beta cells responsible for insulin production, resulting in higher blood glucose levels. ^19^

In a study involved COVID-19 patients confirmed through PCR testing and placed in isolation, revealed that 60% of the patients exhibited elevated blood glucose levels, while 40% did not. These results indicate that more than half of the infected individuals experienced an increase in glucose levels. ^20^ Additionally, it was observed that patients with elevated glucose levels had lower insulin levels compared to those with normal glucose levels, providing support for the theory that COVID-19 may lead to the destruction of pancreatic beta cells and subsequently decrease insulin secretion. These findings align with the study’s conclusion that the virus diminishes insulin secretion by directly impacting beta cells in the pancreas. ^20^

On the other hand, earlier studies indicated that elevated level of cortisol hormone in the blood can directly decrease insulin secretion by impacting the secretion rate of beta cells in the pancreas. This has been observed in laboratory experiments involving cultured beta cells and in experiments with genetically modified mice. These studies revealed that high cortisol levels reduce insulin secretion by inhibiting hormone release. Moreover, research also showed that patients with significantly elevated cortisol levels were treated with dexamethasone, prescribed at a dosage of 4 mg twice daily, along with antibiotics like Sulfalaxin, azithromycin, and amantadine. This treatment approach was initially adopted for all COVID-19-infected patients at the beginning of the pandemic. Dexamethasone, a member of corticosteroids, which are synthetic versions of cortisol used to alleviate inflammation, was given to patients facing respiratory distress and requiring oxygen therapy. However, this medication can result in various complications, such as increased levels of triglycerides, cholesterol, and blood glucose. Therefore, patients taking this synthetic hormone, in addition to the natural cortisol produced in their bodies, which escalates in response to stress, tension, and anxiety (hence, its label as the “stress hormone”), may experience changes that contribute to heightened blood glucose levels known as stress hyperglycemia. ^21^

Several studies have suggested a positive association between CRP and prediabetes, including hyperglycemia and metabolic syndrome. However, these studies have not definitively confirmed whether high blood glucose leads to inflammation and consequently stimulates CRP secretion, or if high CRP leads to an increase in blood glucose through its impact on insulin. ^22^ Additionally, acute inflammation in the liver cells, such as in the case of hepatitis, can cause insulin resistance and subsequent high blood glucose, contributing to the onset of type II diabetes. ^23^ While some studies have demonstrated acute inflammation in liver cells indicated by a clear rise in liver enzymes GOT and GPT, it was noted that there was no evidence of insulin resistance as insulin levels were low, thus excluding hepatitis as the cause of high blood glucose. Consequently, the elevation of liver enzymes can be attributed to an inflammatory condition in the body as a result of viral infection.

Comparing the findings of this study with the existing theories attempting to explain the relationship between the COVID-19 virus and high blood sugar, it is evident that factors leading to elevated blood glucose were present in the patients studied. It can thereby be inferred that these factors collectively contributed to an increase in their blood glucose levels. These factors are likely to produce similar effects in patients with diabetes prior to contracting the COVID-19 virus, possibly leading to a significant rise in blood glucose levels and even resulting in fatalities. Moreover, some studies have indicated that several patients developed diabetes after recovering from COVID-19 infection, implying a direct impact of the virus on beta cells in the pancreas, resulting in reduced insulin secretion, as supported by the findings of this study.

In this study, the insulin secretion of individuals was determined using HOMA-β as a model to study pancreatic β-cell function, The results of this study showed that there was a statistically significant difference of HOMA-β values between individuals with high blood glucose and normal blood glucose. The results showed that β-cell functional decline is associated with low HOMA-β value in individuals with high blood glucose. The study results indicated that the HOMA-IR value was high in individuals with normal blood glucose concentration, and high HOMA-IR means insulin resistance, indicating that they are in the pre-diabetes stage. In this group, the HOMA-B value was high compared to the other with high glucose, and the high value of HOMA-[with the high value of HOMA-IR indicates that the beta cells in the pancreas work hard to secrete more insulin because there is resistance to the effect of insulin. and this process occurs in the pre-diabetes stage, which confirms that this group is at increased risk of developing type 2 diabetes if the underlying causes are not treated. These results consistent with some studies that have indicated that the COVID-19 leads to the destruction of beta cells in the pancreas^7^, which leads to a decrease in insulin concentration and thus an increase in blood glucose concentration.

## Conclusion

Blood glucose level was increased in some patients infected with the COVID-19, accompanied by decrease in the concentration of the hormone insulin they have. Our findings also revealed an increase in the variables that are related to raising the concentration of glucose in the blood, namely cortisol and fats, accompanied by a strong positive correlation with these variables.

## Data Availability

All data produced in the present study are available upon reasonable request to the authors

## Acknowledgments

All the work was self-financing, without assistance from any government agency, and all the researchers participating in this research completely agreed on everything that was conducted during the work period.

